# Atrophy-centered subtyping of mild cognitive impairment

**DOI:** 10.1101/2020.11.28.20238964

**Authors:** Kichang Kwak, Kelly S. Giovanello, Martin Styner, Eran Dayan, for the Alzheimer’s Disease Neuroimaging Initiative

**Affiliations:** Biomedical Research Imaging Center, University of North Carolina at Chapel Hill; Department of Psychology and Neuroscience, University of North Carolina at Chapel Hill; Department of Computer Science, University of North Carolina at Chapel Hill; Department of Psychiatry, University of North Carolina at Chapel Hill; Department of Radiology, University of North Carolina at Chapel Hill

## Abstract

Mild cognitive impairment (MCI) is considered as the transitional phase between normal cognitive aging and Alzheimer’s disease (AD). Nevertheless, trajectories of cognitive decline vary considerably among individuals with MCI. To address this heterogeneity, subtyping approaches have been developed, with the objective of identifying more homogenous subgroups and ultimately improving prognostic outcomes. To date, subtyping of MCI has been based primarily on cognitive performance measures, often resulting in indistinct boundaries between the proposed subgroups and limited validity. The degree to which markers of neurodegeneration such as brain atrophy can be used to subtype MCI into biologically and clinically meaningful subgroups remains unclear. Here we introduce and validate a data-driven subtyping method for MCI based solely upon measures of atrophy derived from structural magnetic resonance imaging (MRI). We trained a dense convolutional neural network to differentiate between patients with AD and age-matched cognitively normal (CN) subjects based on whole brain MRI features. We then deployed the trained model to classify individuals with MCI, as MCI-CN or MCI-AD, based on the degree to which their whole brain gray matter volume resembles CN-like or AD-like patterns. We subsequently validated the model-based subgroups using cognitive, clinical, fluid biomarker, and molecular neuroimaging data. Namely, we observed marked differences between the MCI-CN and MCI-AD groups in baseline and longitudinal cognitive and clinical rating scales, disease-free survival, cerebrospinal fluid (CSF) levels of amyloid beta and tau, fluorodeoxyglucose (FDG) and amyloid PET. Overall, the results suggest that patterns of atrophy in MCI are sufficiently distinct and heterogeneous, and can thus be used to subtype individuals into biologically and clinically meaningful subgroups.

## Introduction

Mild cognitive impairment (MCI) is often construed as the transitional stage between normal aging-related cognitive decline and Alzheimer’s disease (AD) *(1, 2)*. However, MCI is associated with marked etiological heterogeneity *(3)*. While the yearly risk of progression from MCI to AD is set at around 10% to 12% *(4–6)*, not all individuals with MCI eventually progress to AD and many demonstrate different outcomes, including the development of non-AD dementia or other neuropsychiatric conditions *(7, 8)*, or reversion to cognitively normal (CN) status *(9)*. Despite its ubiquity, the heterogeneity of MCI remains poorly understood, challenging further progress in research and care.

Attempts to constrain the heterogeneity of MCI via subtyping approaches have been proposed by several groups *(10–12)* almost exclusively relying on cognitive subtyping, that is, classification into subtypes which is based on subjects’ performance in cognitive tests and tasks *(11, 13)*. For example, a common subtyping framework defines individuals with MCI as amnestic and non-amnestic, depending on whether or not memory loss is a predominant feature *(11)*. Other common subtyping approaches further classified MCI as being either single- or multiple-domain as a function of the number of cognitive domains where decline is observed *(14)*. More recently, studies have shown that a more comprehensive subtyping for MCI can be achieved based on similarities in neuropsychological test scores using clustering techniques *(15, 16)*.

While cognitive subtyping has been instrumental in delineating the various dimensions of cognitive performance that are affected in MCI, cognitive subtypes may suffer from insufficiently distinct boundaries and their validity has been questioned *(17)*. Moreover, a shift in the definition of preclinical, prodromal and clinical AD from a syndromic to a biological construct will hopefully facilitate advances in the identification of new treatment targets and to increased precision in interventional clinical trials *(18)*. Indeed, the recently proposed “AT(N)” framework for AD research *(19, 20)*, attempts to provides accurate, biologically-centered definitions for AD research based on multi-domain biomarkers for β-amyloid deposition (‘A’), pathologic tau (‘T’), and neurodegeneration (‘N’). Nevertheless, biomarkers for neurodegeneration, particularly those based on structural magnetic resonance imaging (MRI), are not specific to AD, and may be attributable to various other comorbidities *(18)*. Consequently, the degree to which heterogeneity in atrophy patterns in MCI can be leveraged to subtype individuals into homogeneous subgroups remain unclear.

In the current study, we tested whether patterns of brain atrophy, derived from MRI, are sufficient in allowing for the subtyping of MCI into biologically and clinically distinct subgroups. We propose and validate a novel data-driven subtyping approach based upon deep learning. We first train a dense convolutional neuronal network (CNN) to differentiate between AD and CN based on whole brain gray matter morphometric data. We then deploy the trained CNN to classify MCI subjects into two subgroups, MCI-AD and MCI-CN, corresponding to MCI subjects with AD-like and CN-like morphometric characteristics, respectively. We also identified the major regional atrophy patterns contributing to the differentiation between the two MCI subtypes through occlusion analysis *(21)*. The resulting labels were then validated against cerebrospinal fluid (CSF) biomarkers for β-amyloid (Aβ) and tau, baseline fluorodeoxyglucose (FDG) and amyloid positron emission tomography (PET), as well as baseline and longitudinal cognitive scores. Finally, we evaluated the degree of overlap between the modeling-based labels and those obtained through cognitive subtyping.

## Results

### Participant Characteristics

We analyzed data from 489 subjects, obtained from the Alzheimer’s Disease Neuroimaging Initiative (ADNI) database. Our proposed modeling approach (see below) utilized AD and CN data for training, and MCI data for testing. Following the proposed NIA-AA guidelines *(18)*, AD subjects were included in the analysis if they displayed abnormal CSF Aβ_42_ and p-tau_181_ levels (denoted henceforth as, A+T+). CN subjects were included in the analysis only if they displayed normal CSF Aβ_42_ and p-tau_181_ levels (henceforth, A−T−). The demographic characteristics of subjects in the AD and CN groups are shown in Table 1. When comparing the AD, CN, and MCI groups (Table S1), there were significant difference in Clinical Dementia Rating (CDR: *F*_*2,596*_ = 458.2, *p* < 0.001), Alzheimer’s Disease Assessment Scale (ADAS: *F*_*2,596*_ = 177.90, *p* < 0.001), and CSF biomarker concentrations (A*β*_42_: *F*_*2,596*_ = 146.45, *p* < 0.001, p-tau_181_: *F*_*2,596*_ = 86.08, *p* < 0.001). Post hoc comparisons with the Tukey test revealed significant pairwise difference (all p’s < 0.001) between the 3 groups in all cognitive measures and CSF biomarker concentrations. No significant differences were observed between the groups in age (*F*_*2,596*_ = 2.16, *p* = 0.12) and gender distribution (*χ*^2^ = 3.06, *p* = 0.22).

**Table 1.** Demographics.

|  | **AD (A+T+)** | **CN (A-T-)** | **MCI** |
| --- | --- | --- | --- |
| N | 110 | 109 | 380 |
| Age | 72.91 (8.03) | 70.98 (5.32) | 72.15 (7.19) |
| Gender, female | 46 (41.82%) | 57 (52.29%) | 166 (43.68%) |
| ADAS | 21.97 (7.03) | 6.97 (3.05) | 9.64 (4.43) |
| CDR | 4.58 (1.72) | 0.0 (0.07) | 1.49 (0.89) |
| $A\beta_{42}$ (pg/ml) | 597.79 (158.44) | 1454.75 (247.64) | 987.22 (425.97) |
| p-tau<sub>181</sub> (pg/ml) | 40.41 (14.97) | 16.4 (2.82) | 27.59 (14.91) |

Continuous variables are presented as means, with SDs and categorical variables are presented as % in parentheses. Abbreviations: AD=Alzheimer's disease, CN=cognitively normal, MCI=mild cognitive impairment. N=number of subjects, ADAS=Alzheimer's disease assessment scale, CDR=clinical dementia rating, $A\beta_{42}$ =beta-amyloid42, p-tau<sub>181</sub>=phosphorylated-tau181, SD=standard deviation.

### A deep learning model for subtyping MCI subjects

Our major objective in the current study was to develop a deep learning modeling framework for subtyping MCI. To that end we utilized a dense CNN architecture (Fig. 1) *(22)*, which relies on whole brain gray matter (GM) density as input data. The model was first trained to differentiate AD and CN, with the assumption that these 2 distinct groups would provide the model with an adequate distribution of morphometric features sufficient for subtyping MCI subjects. Data augmentation was applied within the training data to increase its size and improve the performance of the model and its generalizability. We used 5-fold cross-validation within the training set to optimize and fine-tune the model’s performance, finding similar performance across the different folds (Fig. S1). The model with the best performance achieved maximal accuracy of 93.75%, with an area under the curve (AUC) of the receiver operating characteristic (ROC) of 0.983 (Fig. S1). We subsequently deployed the model on the MCI data (Fig. 1), formalized as a binary classification problem with class labels AD and CN. Then, output values less than the default threshold of 0.5 were assigned to the class CN (henceforth, MCI-CN) and values greater than or equal to 0.5 were assigned to the class AD (henceforth, MCI-AD). We also validated the group differences across various thresholds for binary classification and observed relatively little differences as a function of threshold (Fig. S2).

**Fig. 1.**
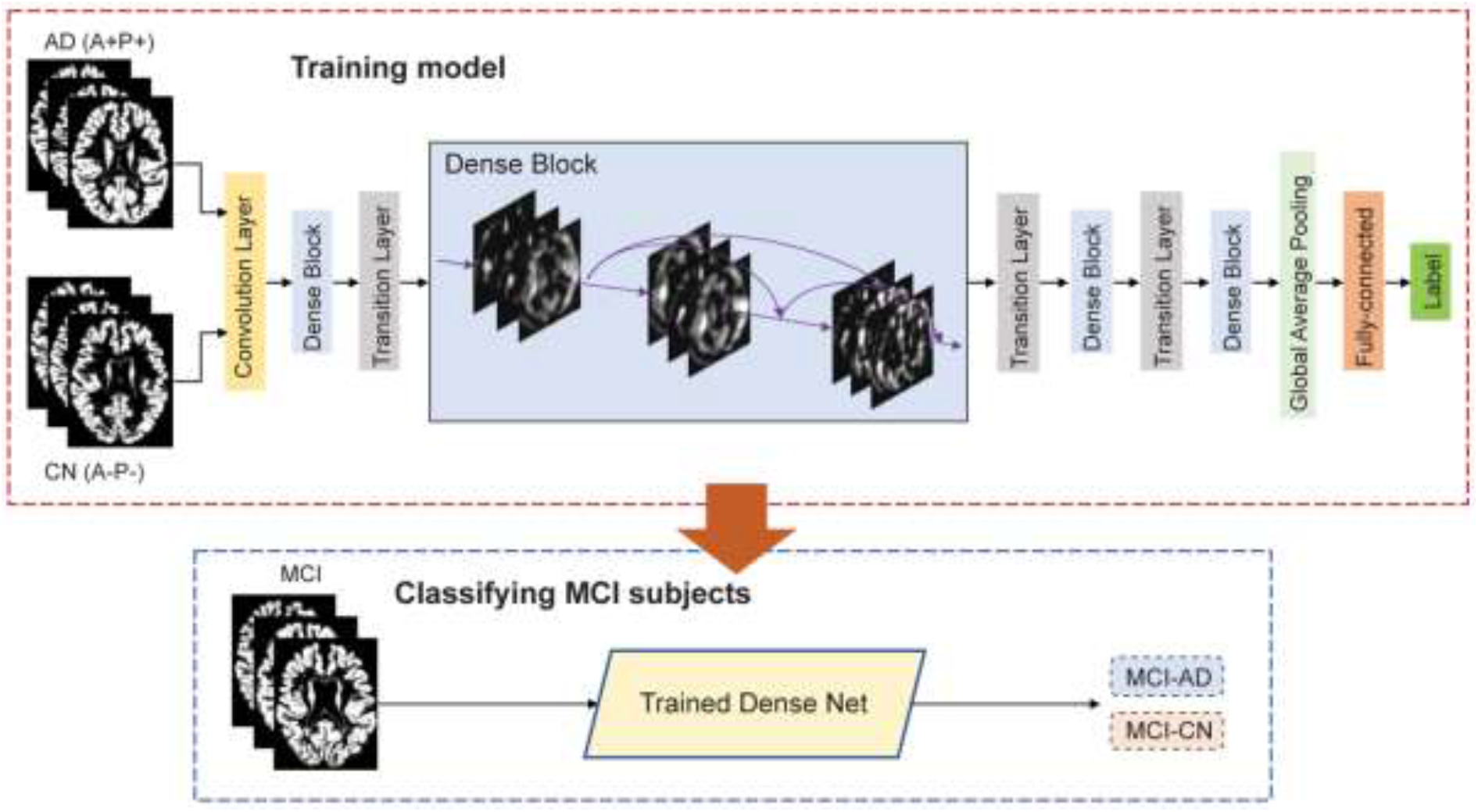
Study methods. Illustration of proposed deep learning framework. A dense convolutional neural network is trained to differentiate patients with Alzheimer’s disease (AD) and cognitively normal (CN) controls based on whole brain gray matter morphometric data. Subsequently, the trained model is deployed to classify individuals with mild cognitive impairment (MCI), into two groups, MCI-AD and MCI-CN based on structural morphometric data.

### Validation of model-based MCI subtypes with CSF biomarker concentrations and cognitive scores

Our data-driven approach for subtyping MCI resulted in two subgroups, MCI-AD and MCI-CN. We next assessed the validity of these two data-driven labels. We first compared the prevalence of the various biomarker profiles *(18)* in each of the subgroups, based on each subject’s CSF Aβ_42_ and p-tau_181_ biomarkers. Each subject was rated as either positive (i.e., abnormal) or negative (i.e., normal) in each biomarker, based on previously published cut-off values *(18)* (See Fig. S3). This resulted in 4 different profiles (A+T+, A+T−, A−T+, and A−T−, where ‘A’ denotes Aβ, and ‘T’ denotes tau) (Fig. 2A, B). The prevalence of the biomarker profiles differed significantly between the MCI-AD and MCI-CN subgroups (*χ*^2^ = 21.40, *p* < 0.001). In particular, subjects with an abnormal CSF Aβ_42_ or p-tau_181_ were more common in the MCI-AD (CSF Aβ_42_: 71.6%; p-tau_181_: 69.3%) than in the MCI-CN group (CSF Aβ_42_: 50.6%; p-tau_181_: 50.2%). Subjects with an abnormal CSF Aβ_42_ and p-tau_181_ were more prominent in the MCI-AD (A+T+: 55.1%) than in the MCI-CN group (A+T+: 32.8%). In contrast, subjects with a normal CSF Aβ_42_ and p-tau_181_ were more common in the MCI-CN (A−T−: 32.0%) than in MCI-AD group (A−T−: 14.2%).

**Fig. 2.**
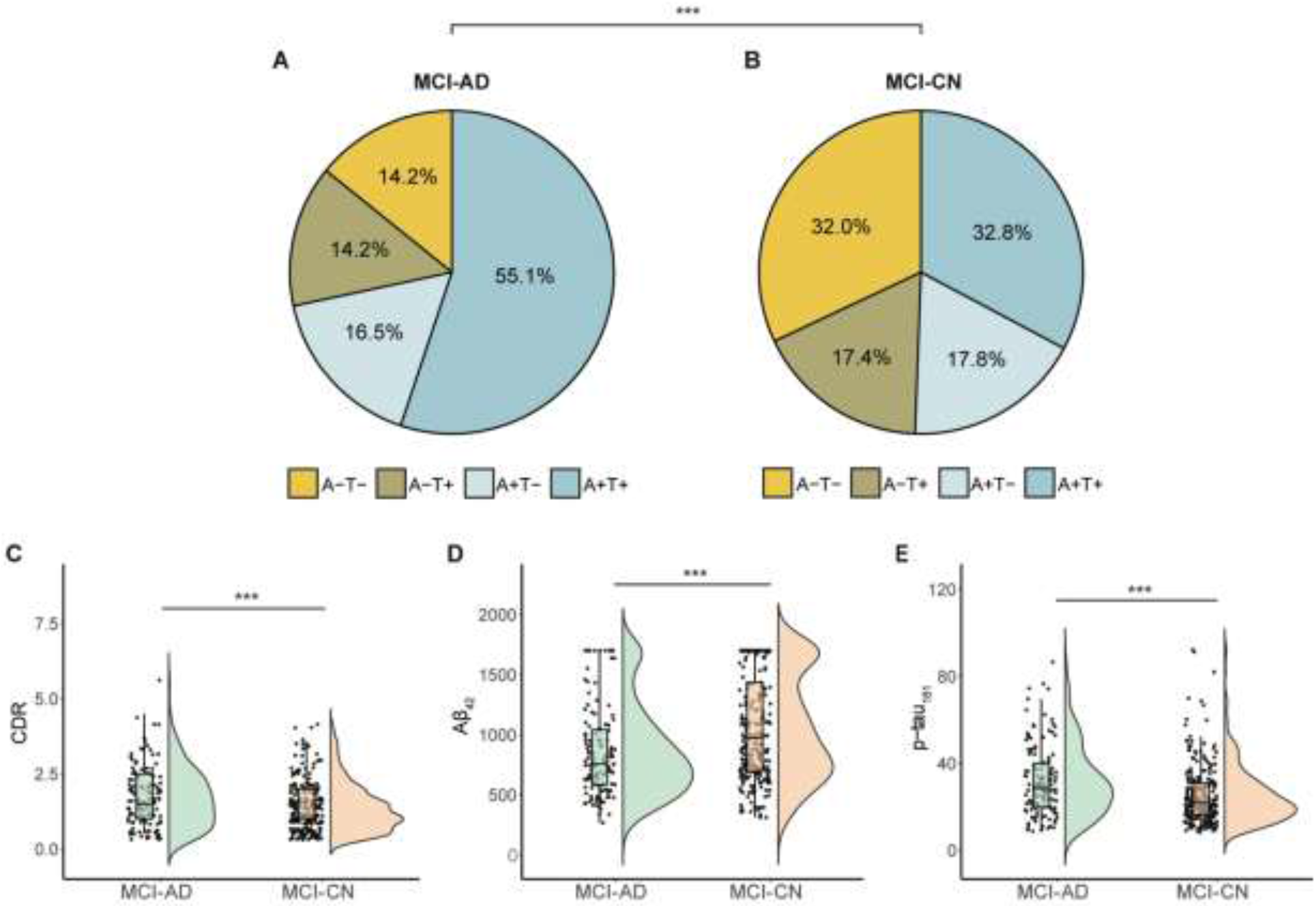
Validation of the MCI subgroups using fluid biomarker and cognitive data. Subjects in the MCI-AD and MCI-CN groups were rated as Amyloid (A) and p-tau (T), positive or negative, based on the CSF A*β*_42_, and p-tau_181_ biomarkers. Pie charts depict the biomarker score combinations in the MCI-AD (**A**) and MCI-CN (**B**) subgroups. These score distributions were significantly different between the two subgroups. Box plots show differences in CDR scores (**C**), CSF A*β*_42_ (**D**), and CSF p-tau_181_ (**E**). Abbreviations: AD=Alzheimer’s disease, CN=cognitively normal, MCI=mild cognitive impairment, CDR=clinical dementia rating. \*\*\**p*<0.001.

We then investigated baseline differences in demographic characteristics, cognitive scores and continuous CSF concentrations between the MCI-AD and MCI-CN subgroups. There was a significant difference between the two subgroups in age (*t*_*378*_ = 6.44, *p* < 0.001), but not in gender distribution (*χ*^2^ = 3.49, *p* = 0.06). Comparing cognitive scores between the MCI-AD and MCI-CN subgroups reveled significant differences in CDR (*t*_*378*_ = 3.46, *p* < 0.001; Fig 2C) and ADAS (*t*_*378*_ = 6.51, *p* < 0.001; See Fig S4) scores. All significant results were retained when controlling for age (all p values < 0.01). The comparison of CSF concentrations between the MCI-AD and MCI-CN subgroups revealed significant differences in CSF Aβ_42_ (*t*_*378*_ = 4.55, *p* < 0.001; Fig 2D) and in p-tau_181_ (*t*_*378*_ = 3.81, *p* < 0.001; Fig 2E). The significant results were retained when controlling for age (both p values < 0.001).

### Comparison of PET uptake in subtyped MCI

We next assessed group differences in PET uptake, focusing on Aβ- and FDG-PET. While Aβ-PET and CSF measures of Aβ are generally properly correlated with one another, a certain degree of discordance between the two markers has been consistently reported *(23, 24)*. Our analysis above revealed that the MCI-AD and MCI-CN subgroups differed in CSF Aβ_42_ levels. We thus next aimed to complement this analysis by also comparing the subgroups across Aβ-PET. There were significant differences between the two subgroups in Aβ-PET (*t*_*377*_ = 4.36, *p* < 0.001; Fig 3A), with lower levels observed in the MCI-AD group, relative to the MCI-CN group. We have additionally examined group differences in FDG-PET uptake. Metabolic imaging studies utilizing FDG-PET for AD diagnosis are common *(25)*, and this modality has been proposed as a reliable and valid marker for neurodegeneration in AD *(18)*. We observed significant group differences in FDG-PET (*t*_*377*_ = 5.53, *p* < 0.001; Fig 3A), with lower uptake values obtained in the MCI-AD group. Group differences in both Aβ-PET and FDG-PET were retained after adjusting for the effect of age. We have additionally assessed group differences in PET uptake, comparing normal/abnormal uptake values after binarizing the data with established cut-off values *(26, 27)*. The prevalence of normal and abnormal Aβ-PET and FDG-PET differed between the two subtyped subgroups (Fig. S5). These differences were apparent in both Aβ-PET and FDG-PET (Aβ-PET: *χ*^2^ = 12.35, *p* < 0.001; FDG-PET: *χ*^2^ = 14.70, *p* < 0.001). In particular, there were more subjects with abnormal Aβ-PET in the MCI-AD (Aβ-PET+: 71.4%) than in MCI-CN group (Aβ-PET+: 52.6%). Similarly, abnormal FDG-PET was more common in the MCI-AD (FDG-PET+: 48.4%) than in MCI-CN group (FDG-PET+: 28.5%).

**Fig. 3.**
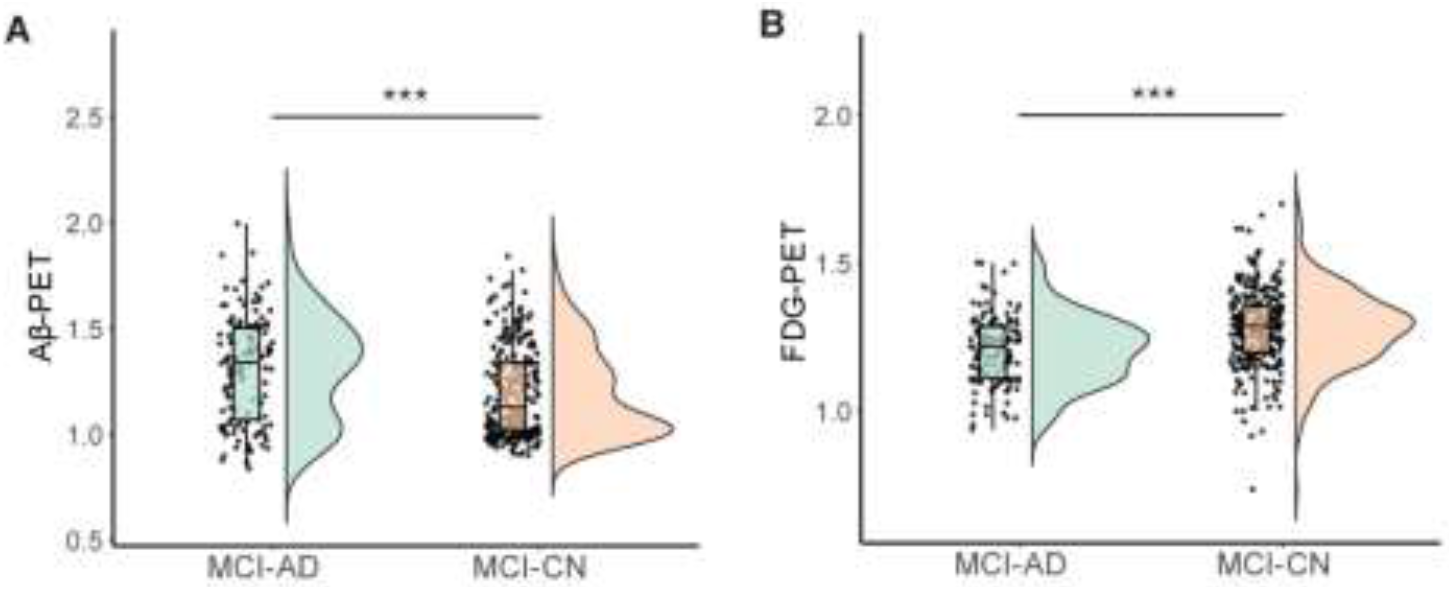
Comparison of the MCI subgroups using PET uptake data. Boxplots show the comparison of A *β* -PET (**A**) and FDG-PET (**B**) uptake between the MCI-CN and MCI-CD subgroups. In both measurements, group differences were statistically significant. Abbreviations: AD=Alzheimer’s disease, CN=cognitively normal, MCI=mild cognitive impairment, A *β* -PET=beta-amyloid positron emission tomography, FDG-PET=fluorodeoxyglucose-positron emission tomography. \*\*\**p*<0.001

### Contribution of brain regions in subtyped MCI: occlusion analysis

The results suggest that patterns of whole brain gray matter are sufficient for differentiating MCI subjects into 2 distinct subgroups. As our approach utilizes whole brain gray matter features, the major regional contributors to the model’s output remain unclear. We thus next examined the relative lobar (frontal, parietal, medial temporal, lateral temporal, occipital, and cingulate) contribution to the performance of the model, through occlusion analysis, as proposed previously *(21)*. Briefly, we retested the deep learning model iteratively, occluding a bilateral binary mask composed of each lobe from the model’s test-set input data (Fig. 4A). This was achieved by setting the intensity values of each lobe to zero on each iteration. The Percentages of change in the model’s output, with respect to the original results for classifying MCI subgroups were ranked and then compared across the different occluded lobes (See Fig. 4B). We found that the occlusion of the medial temporal and lateral temporal lobes led to dramatic changes in the model’s output (Fig. 4C), relative to the original results. On the other hand, occlusion of the occipital and cingulate lobes had relatively little effect, resulting in model output that resembled the original results (Fig. 4C). Thus, the medial and lateral temporal lobes had the largest impact on the performance of the model and on the classification of MCI subjects into two distinct subgroups.

**Fig. 4.**
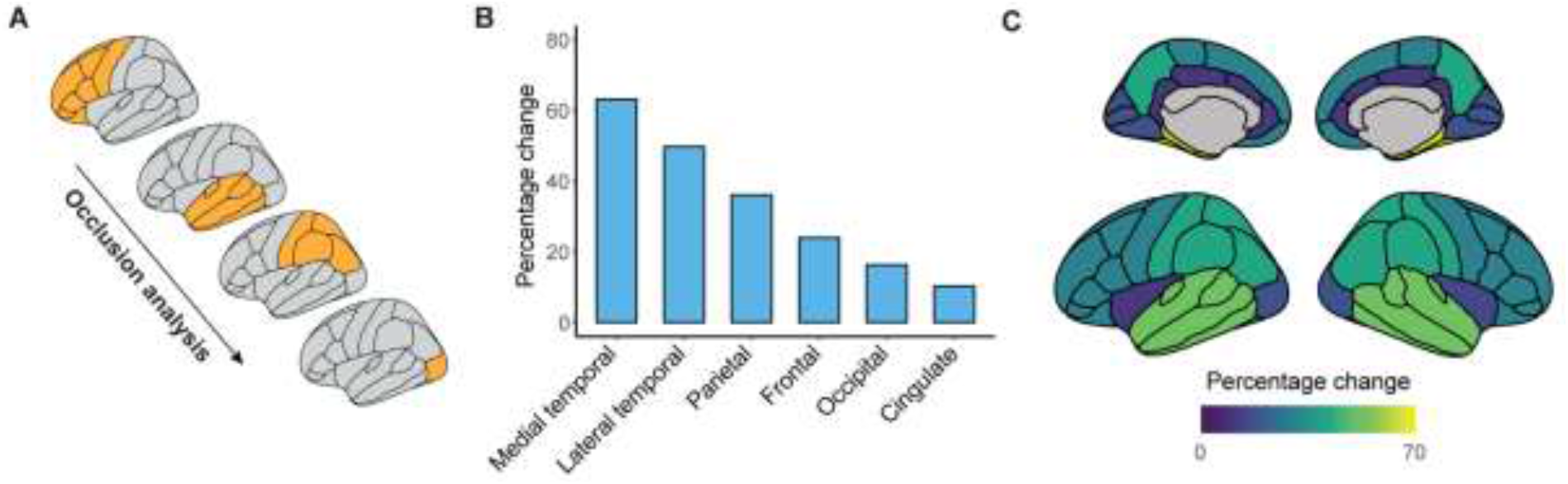
Identifying the major contributors to atrophy-centered subtyping of MCI via occlusion analysis. (**A**) Schematic illustration of the occlusion analysis. The testing phase in the deep learning model was repeated, whereby in each step, cortical lobes were occluded from the input data (temporal lobes were further divided to medial and lateral). Percentage of change with respect to the original results for classifying MCI subgroups were ranked. (**B**) The results of the occlusion analysis are shown, in each tested lobe. Shown are percentages of change with respect to the original intact model. (**C**) Percentage of change following occlusion analysis, in each cortical lobe, superimposed on medial and lateral cortical surface models. Abbreviations: MCI=mild cognitive impairment

### Longitudinal analysis of cognitive changes in the MCI subgroups

Our analysis reveals marked baseline differences between the MCI-AD and MCI-CN subgroups. These observed differences, nevertheless, cannot be taken to imply that the two MCI subgroups also differ in their prognostic outcomes. We next set out to evaluate if the MCI-AD and MCI-CN subgroups also exhibit differences in the progression to AD and in longitudinal cognitive performance. This analysis focused on subjects with data from at least 3 follow-up visits. Individual trajectories of longitudinal changes in cognitive performance varied between the two MCI subgroups (Fig. S6). Survival analysis *(28)* revealed marked differences between the two subgroups in their progression to AD (Log-rank test; *χ*^2^ = 64.40, *p* < 0.001), with the MCI-AD subgroup showing faster progression, relative to the MCI-CN subgroup, where slower progression was observed over time (Fig. 5A). We next used repeated measures-analysis of variance (RM-ANOVA) to compare changes in cognitive performance from baseline to the 2^nd^ year follow-up visit, with group (MCI-AD, MCI-CN) as the between-subjects factor and time (baseline, follow-up) as the within-subject, repeated-measure factor. Focusing on CDR scores (Fig. 5B), this analysis revealed a significant interaction (group × time; *F*_*1,358*_ = 14.92, *p* < 0.001), along with significant main effects for group (*F*_*1,358*_ = 27.87, *p* < 0.001) and time (*F*_*1,358*_ = 38.91, *p* < 0.001). Thus, the MCI-AD group showed more pronounced changes in CDR scores between the testing sessions. Similarly, in the analysis of ADAS scores (See Fig. S7), a significant interaction (group × time; *F*_*1,358*_ = 7.56, *p* < 0.001) was observed, along with significant main effects for group (*F*_*1,358*_ = 5.92, *p* = 0.02), and time (*F*_*1,358*_ = 87.12, *p* < 0.001).

**Fig. 5.**
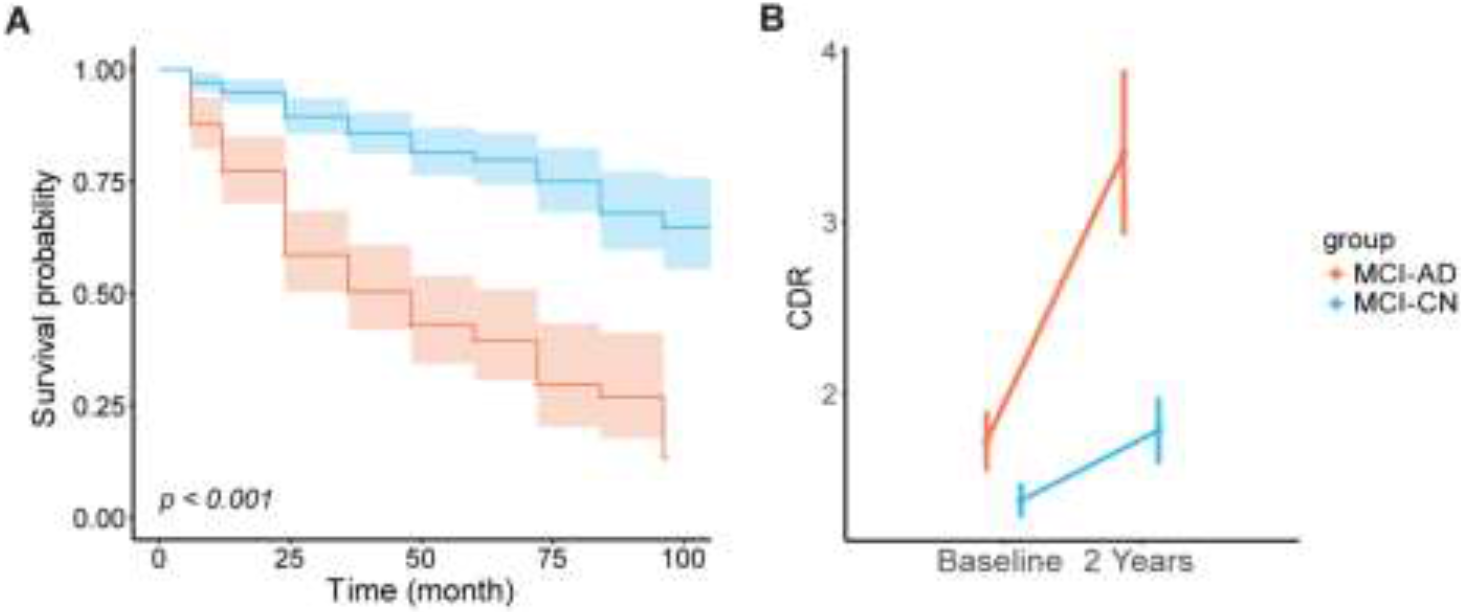
Longitudinal comparison of the MCI subgroups. (**A**) Kaplan-Meier plots depicting disease-free survival in the MCI-AD and MCI-CN subgroups. The MCI-CN group showed significantly better disease-free survival over time (log-rank test). Shaded areas depict confidence intervals (**B**) Longitudinal changes in CDR scores, displayed by the two MCI subgroups, tested with a RM-ANOVA. Abbreviations: AD=Alzheimer’s disease, CN=cognitively normal, MCI=mild cognitive impairment, CDR=clinical dementia rating, RM-ANOVA=repeated measures-analysis of variance.

### Concordance between the current MCI subtyping approach and cognitive subtyping

The data-driven approach proposed here subtypes MCI subjects solely based on patterns of whole brain gray matter volume. As the vast majority of existing subtyping approaches for MCI are based on cognitive profiles *(17)*, we wished to determine the extent of concordance between the current MCI subtyping approach and cognitive subtyping. We focused the comparison on a recently-introduced subtyping method where neuropsychological assessments are clustered into distinct subgroups *(15)*. We first clustered subjects (n=374 subjects who had available data) based on their neuropsychological assessments (Table S2). This resulted in 4 subgroups: *Dysnomic* (37.43% of subjects), *amnestic* MCI (36.63), *Dysexecutive* (6.95%), and *Cluster-Derived Normal* (18.98%) (Fig. 6A). These 4 subgroups showed significant group differences in the 6 neuropsychological assessments used for clustering (*p* < 0.001). Post-hoc comparisons with the Tukey multiple comparison test revealed that the *Dysnomic* group performed worse than all other groups in 5/6 measures of language. The *Dysexecutive* group performed worse than all other groups in assessments of attention/executive function, and the amnestic MCI group performed worse than the *Dysnomic* and *Cluster-Derived Normal* groups in assessments of memory function. Thus, the clustering of neuropsychological assessments resulted in distinct MCI subtypes, as previously reported *(15)*. We next compared the distributions of the neuropsychological subtypes within the MCI-AD and MCI-CN subgroups. We found significant differences between the two distributions (*χ*^2^ = 30.45, *p* < 0.001), observing more *Dysnomic* and *Dysexecutive* subjects in the MCI-AD (60%) than in the MCI-CN group (36.5%). On the other hand, as expected, the *Cluster-Derived Normal* profile, which shows the normal range of cores across all neuropsychological measures, was more common in the MCI-CN (24.9%) than in the MCI-AD group (7.2%). Interestingly, the prevalence of the Amnestic MCI profile was similar between the MCI-AD (32.8%) and MCI-CN subgroups (38.6%). Altogether, while both the MCI-AD and MCI-CN subgroups displayed impairments in memory function, the former subgroup displayed more significant deficits in attention/executive and language function than the latter subgroup.

**Fig. 6.**
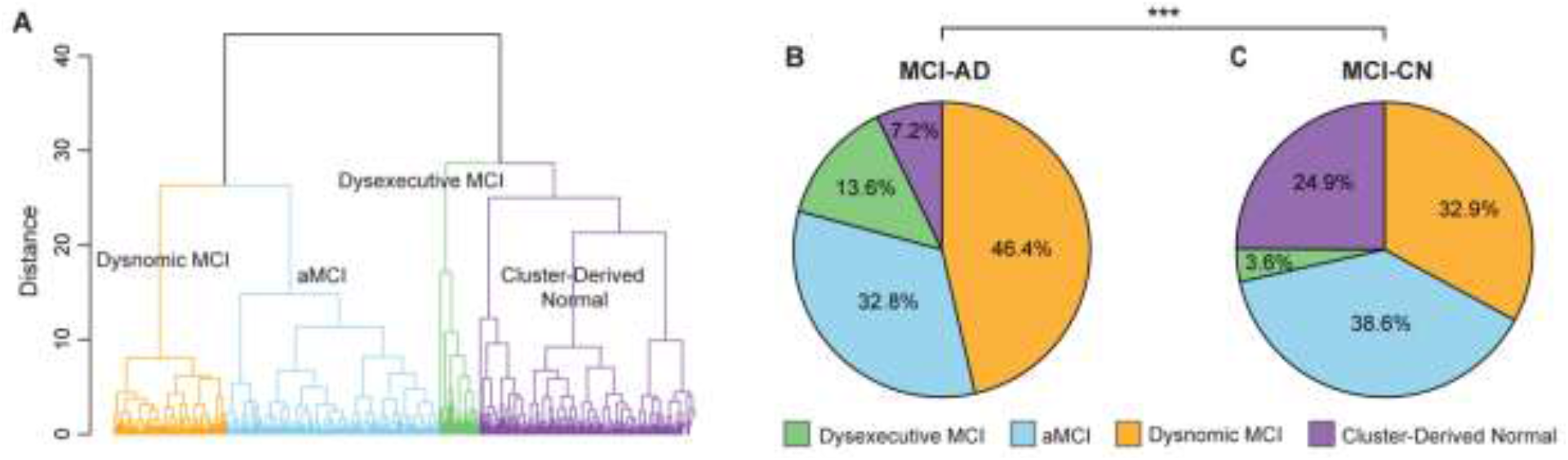
Overlap between atrophy-centered and neuropsychological subtypes. (**A**) Hierarchical clustering on neuropsychological data was used to define 4 MCI subtypes within the dataset used here (dysnomic MCI, aMCI, dysexecutive MCI, and cluster-derived normal). Pie charts show the prevalence of these subtypes in the MCI-AD (**B**) and MCI-CN (**C**) groups. The two distributions were significantly different. Abbreviations: AD=Alzheimer’s disease, CN=cognitively normal, MCI=mild cognitive impairment, aMCI=amnestic mild cognitive impairment. \*\*\**p*<0.001

## Discussion

Subtyping approaches for MCI have been proposed as a remedy for the large etiological heterogeneity characteristic of this elusive stage in cognitive aging. Current subtyping approaches primarily rely on neuropsychological profiles and may often result in blurred boundaries between subgroups and limited validity *(17)*. Here, we propose a novel data-driven subtyping approach, which utilizes CNNs to divide MCI subjects into subgroups based on the extent to which their brain atrophy patterns resemble those observed in AD as opposed to CN data. This approach resulted in two subgroups, MCI-AD and MCI-CN, denoting closer correspondence in gray matter patterns with the AD and CN, respectively. We then comprehensibly validated the model-based subgroups, findings marked group differences in baseline CSF biomarker concentrations and PET uptake, along with baseline and longitudinal cognitive performance scores. Through occlusion analysis *(21)* we investigated lobar contribution to the performance of the deep learning model, reporting that it mostly relied on gray matter volume from the medial and lateral temporal lobes. Finally, we found a limited degree of overlap between the current subtyping approach and that based on neuropsychological examination.

The purpose of the current study was to test whether distinct subgroups with differing structural brain atrophy patterns could be delineated within a heterogeneous clinical sample of individuals diagnosed with MCI. To that end, we utilized a deep learning framework, rather than other machine learning models, such as support vector machine, primarily since there has been a growing body of research demonstrating the utility of deep learning models based on MRI-derived features in various tasks, such as diagnostic prediction *(29)*, image reconstruction *(30)* and segmentation *(31)*, and prognostic prediction of disease progression *(32)*. Our choice to utilize a deep learning framework was further motivated by the assumption that complex and non-linear relationships exist between whole brain structure and progression of MCI/AD. Similar to other machine and deep learning models where “transfer learning” *(33)* is applied, we propose a classification framework which is trained on a domain different than the one being tested *(34–36)*. However, rather than evaluating the performance of the model against clinically-defined labels (e.g., progressive and stable MCI, or AD convertors and non-convertors), our approach was to re-label data from individuals with MCI based on its proximity to the model’s trained labels, that is, AD and CN.

We found robust differences between the MCI-AD and MCI-CN subgroups in CSF biomarker concentrations, cognitive scores, and PET uptake, suggesting our data-driven method subtypes MCI into biologically and clinically distinct subgroups. Moreover, the prevalence of biomarker profiles, defined based on established cut-off thresholds for the CSF Aβ_42_ and p-tau_181_ biomarkers differed significantly between the MCI-AD and MCI-CN subgroups. Namely, abnormal CSF Aβ_42_ (A+) and p-tau_181_ (T+) were more prevalent in the MCI-AD group. These findings are consistent with earlier studies where it was shown that positive CSF biomarker concentrations (AD-pathological) can predict conversion from MCI to AD with accuracy larger than 80% *(37, 38)*. We additionally found more pronounced cognitive impairment, as assessed with the CDR and ADAS scores, in the MCI-AD subgroup, relative to the MCI-CN subgroup. The two subtyped subgroups also exhibited marked differences in Aβ-PET, a marker of amyloid deposition, and FDG-PET, a commonly-used marker of neurodegeneration *(18)*. The topographical distribution of Aβ deposition, assessed with PET is predicative of progression of individuals with MCI to AD *(39)*, with abnormalities appearing long before the onset of clinical symptoms *(40)*. The Aβ-PET abnormalities observed in the MCI-AD subgroup, where longitudinal cognitive outcomes were poorer and progression to AD was quicker, are consistent with these findings. Similarly, the finding that negative FDG-PET was more predominant in the MCI-CN subgroup is consistent with the observation that negative FDG-PET is highly predictive of clinically stable MCI *(41)*.

We complemented the initial analysis, which relied on whole brain gray matter patterns, with an analysis that combined deep learning classification with occlusion analysis *(21)*. This allowed us to identify the major lobar contributors to the performance of the subtyping model. The results revealed that the medial temporal, lateral temporal, and to a lesser extent the parietal lobe were more central to the model’s performance than other lobes. These findings are consistent with earlier studies on AD pathology, where atrophy was reported in the hippocampus, amygdala, and entorhinal cortex *(42, 43)*. Our results also suggest that the occipital lobe played a more minor role in the performance of the model, consistent with Braak’s staging scheme *(44)*, where the occipital lobe is shown to be affected only at later stages of AD *(45)*. Although our results highlight the central role of the medial and lateral temporal lobes in the subtyping of MCI, further examination into the possible involvement of other cortical and subcortical regions is warranted.

We examined the agreement between our atrophy-centered data-driven subtyping approach and that obtained based on neuropsychological assessments. The latter approach groups subjects based on the similarity of their neuropsychological assessments using clustering techniques, and thus goes beyond the more traditional *amnestic*/*non-amnestic* MCI subgrouping studied extensively in the literature *(46)*. However, neuropsychological and biomarker profiles are strongly heterogeneous in subjects who can be classified as having *amnestic* MCI *(47)*, and empirically-derived subtyping approaches depict heterogeneity that is not captured by conventional criteria. *(15)*. In our analysis, we found that the *Cluster-Derived normal* group, where cognitive function is unimpaired, was more predominant in the MCI-CN than the MCI-AD subgroups. (see Fig. 6). Moreover, while the *Amnestic* MCI profile showed a similar distribution in the two subgroups, *dysnomic* or *dysexecutive* subtypes were primarily represented in the MCI-AD subgroup. Overall, our findings demonstrate that the matching between the two subtyping approaches is incomplete. Future research could attempt to combine the two approaches, to achieve neuropsychologically distinct subgroups, that show differing patterns of brain atrophy.

Several limitations should be noted when considering the current results. First, in this proof-of-concept stage we used atrophy patterns to subtype MCI into two distinct subgroups. We acknowledge that a larger number of subgroups would be needed to better capture the heterogeneity of MCI. In principle, our approach could be modified to output more than two subgroups, however, we believe that a larger number of MCI subjects than that used here would be needed in order to achieve robust and generalizable results. Second, as also noted above, we did not attempt to combine neuropsychological and atrophy-centered features in the current study, primarily as our major motivation was to examine if the latter type of features is sufficient in subtyping MCI. Future research could extend our approach to test if a combination of cognitive and neurobiological features better captures the heterogeneity of MCI.

In conclusion, the current study demonstrates that patterns of gray matter atrophy are sufficient for subtyping MCI into biologically and clinically distinct subgroups. These results further highlight the need to consider the heterogeneity of MCI when attempting to understand the pathological mechanisms of dementia, while providing a potential tool for individualized disease prognosis.

## Materials and Methods

### Subjects

Data used in the preparation of this study were obtained from the ADNI. The ADNI was launched in 2003 as a public-private partnership, led by Principal Investigator Michael W. Weiner, MD. The primary goal of ADNI has been to test whether serial MRI, other biological markers, and clinical and neuropsychological assessment can be combined to measure the progression of MCI and early AD. For up-to-date information, see http://www.adni-info.org/. We used AD (n=110) and CN (n=109) data to train the proposed deep learning model. Then, we subtyped MCI (n=380) into subgroups, validating the model’s output with cognitive scores, CSF biomarker levels and PET uptake available in the same subjects. One subject had no FDG-PET uptake data, while 20 subjects were excluded from the longitudinal analysis of cognitive performance since they had data in less than 3 testing time points. Missing data at the 2nd year follow-up was imputed using linear interpolation/extrapolation. All subjects provided written informed consent and the study’s protocol was approved by the local Institutional Review Boards.

### Study design

In this study, we subtyped MCI subjects using a deep learning approach, based on patterns of brain structural atrophy, derived from MRI. We then validated the model’s output (i.e., subgroups) using CSF biomarker, PET and cognitive/clinical data. Specifically, we trained a dense CNN *(48)*, to differentiate data from AD and CN subjects based on whole brain atrophy patterns. To provide the model with adequate well-defined training data, all AD subjects where A+ and T+, that is, they had abnormal levels of CSF A*β*_42_ and p-tau181. CN subjects were all A- and T-, in other words, they showed normal levels in the same CSF biomarkers. Previously determined cutoff values *(18)* for abnormal Aβ42 (Aβ42 < 976.6pg/ml) and p-tau181 (p-tau181 > 21.8pg/ml) were used. We reasoned that training the model with well-differentiated AD (A+ T+) and CN (A- T-) data would allow the model to learn a more discriminative set of pathological features. We then deployed the trained CNN to classify MCI subjects (n=380), as either AD-like or CN-like. We subsequently validated the model’s output labels with baseline molecular and metabolic neuroimaging, cognitive scales and tests and CSF biomarkers. We additionally examined longitudinal changes in cognitive scores (between baseline and the 2^nd^ year follow-up visit, which was the latest visit where imputation for missing data could be achieved reliably) and calculated disease-free survival in the MCI-AD and MCI-CN subgroups, to assess differences in the progression to AD. Finally, we identified the major regions (lobes) contributing to the performance of the model through occlusion analysis *(21)* (see below), and assessed the intersection between the modeling-based labels and those obtained through cognitive subtyping.

### Image acquisition

Structural MRI (T1) data used in the deep learning model were acquired at ADNI sites using 3T scanners and were based on either an inversion recovery-fast spoiled gradient recalled (IR-SPGR) or magnetization-prepared rapid gradient-echo (MP-RAGE) sequences *(49)*. Aβ-PET data were acquired in a 20-min dynamic emission scan, composed of four 5-min frames. The data was acquired 50–70 min after the injection of 10.0 mCi of [^18^F]-AV45. FDG-PET data were acquired in a 30-min dynamic emission scan, composed of six 5-min frames. The data was acquired 30–60 min after the injection of 5.0 mCi of [^18^F]-FDG. PET data ran through a strict quality control procedure to assess image quality. Standardized image preprocessing correction steps were applied to produce uniform data across the ADNI PET cores. These steps included frame co-registration, averaging across the dynamic range, and standardization with respect to orientation, voxel size, and intensity. Full details of the T1 and PET acquisition parameters and imaging processing steps are listed on the ADNI website (http://adni.loni.usc.edu/methods/).

### CSF collection

CSF collection, shipping, aliquoting, storage as well as analysis followed ADNI’s standardized procedures (http://www.adni-info.org/) *(50, 51)*. Collected CSF samples were frozen on dry ice right after collection (within 1 hour). The samples were then shipped overnight, also on dry ice, to the ADNI Biomarker Core laboratory at the University of Pennsylvania. Aliquots (0.5mL) were prepared from the CSF samples and were then stored at −80°C in barcode-labeled polypropylene vials. Samples for Aβ_42_ and p-tau_181_ were then measured using Elecsys immunoassays. The lower and upper technical limits for the Elecsys Aβ_42_ CSF immunoassay were 200 to 1700pg/mL. The limits for the Elecsys p-tau_181_ CSF immunoassay were 8 to 120pg/mL

### Image processing

The deep learning model used for subtyping MCI data utilized whole brain gray matter data. MRI data were analyzed using Statistical Parametric Mapping 12 (SPM12; Wellcome Department of Imaging Neuroscience, Institute of Neurology, London, UK; http://www.fil.ion.ucl.ac.uk/spm) running on MATLAB 9.8.0 (Math-Works, Natick, MA, USA). Briefly, all MR images were aligned to the anterior commissure and segmented into gray matter (GM), white matter and CSF using the unified segmentation procedure *(52)*, implemented in SPM12. To improve the registration of the GM maps, we used the diffeomorphic anatomic registration through an exponentiated lie algebra algorithm (DARTEL) *(53)*. This resulted in more precise spatial normalization to the template. The DARTEL used subject-specific deformation fields to warp the GM map into subject-specific space, resampled at 2mm isotropic voxels. Then the warped GM maps were affine transformed into Montreal Neurological Institute (MNI) space. In addition to using whole brain GM volume data, we evaluated lobar contribution to the performance of the deep learning model through occlusion analysis (see below).

PET imaging data was used to validate the MCI subtypes outputted by the deep learning model. Detailed acquisition and standardized pre-processing procedures used with the PET images are available at the ADNI website (http://adni.loni.usc.edu/methods/). Amyloid PET uptake was calculated by averaging across 4 cortical regions (frontral, anterior cingulate, precuneus, and parietal cortex) relative to the whole cerebellum region *(26)*. Similarly, FDG-PET uptake was calculated by averaging across a set of pre-defined regions (angular gyrus, posterior cingulate, inferior temporal gyrus) relative to pons/vermis reference regions *(57)*.

### Deep learning model architecture

Model architecture: The deep learning model used for subtyping of MCI (See Fig. 1) was based on the DenseNet architecture *(22)*. It consisted of a convolutional layer, 4 dense blocks, 3 transition layers, a global averaging pooling layer and a fully-connected layer. This state-of-the-art convolutional neural network architecture was chosen as it shows excellent feature propagation and classification performance while alleviating the vanishing gradient problem and significantly reducing the number of parameters used by the model *(22)*. First, the whole brain image with dimensions of 91×109×91 was passed through a stack of convolutional layers, where the filters were of size 5×5×5. The convolution stride was set to 1 voxel, while the size of the max-pooling layer was 2×2×2, with a kernel size set at 2×2×2. The dense block consisted of multiple convolution units, which were equipped with a batch normalization layer, leaky rectified linear unit, a 1×1×1 convolutional layer, a 5×5×5 convolutional layer and a dropout layer. Every convolutional unit was connected to all the previous layers via shortcut connections. Dimensionality reduction of feature maps between dense blocks was achieved through the transition layer. The transition layer included a batch normalization layer, a leaky rectified linear unit, a convolutional layer of size 1×1×1, and an averaging pooling layer of size 2×2×2. The global averaging pooling layer was then concatenated and connected through a fully-connected layer. The model’s output values were processed by the fully-connected layer which used a sigmoid activation function. The output layer mapped all values greater than 0.5 as 1 (positive class: MCI-AD) and all values less than or equal to 0.5 as 0 (negative class: MCI-CN), while testing MCI data based on the pre-trained model (Fig. 1).

### Implementation

The Keras application programming interface in TensorFlow 2.0 was used for building the deep learning model. Model training and testing were performed in a parallelized manner with an Ubuntu 18.04.3 operating system, utilizing two Nvidia Tesla V100 graphic cards with 16GB memory each. The model was trained with a mini-batch size of 24 and 200 epochs, and optimized using stochastic gradient descent based on adaptive estimation of first- and second-order moments *(58)* and an exponentially decaying learning rate. The initial learning rate was set at 0.0001 and decayed by 0.9 after every 10000 steps. A dropout layer was added to the dense block, with the dropout rate set to 0.2. In the batch normalization step, beta and gamma weights were initialized with L2 regularization set at 1×10^−4^ and epsilon set to 1.1×10^−5^. In the fully-connected layer, the L2 regularization penalty coefficient was set at 0.01. We observed stability in the model after an iteration of 150 epochs.

### Occlusion analysis

To identify the more specific regional contribution to the performance of the deep learning model and the differentiation between the two MCI subgroups, we integrated occlusion analysis (e.g., *(59)*) into the classification framework. Cortical regions were first segmented with an automated segmentation tool available in FreeSurfer v6.0 (https://surfer.nmr.mgh.harvard.edu/), resulting in a parcellation of the cerebral cortex into 34 sulcul and gyral regions of interest (ROI) per hemisphere, according to the Desikan-Killiany protocol *(54, 55)*. We then merged individual ROIs to 6 lobes: frontal, parietal, medial temporal, lateral temporal, occipital, and cingulate *(56)*. These ROIs were then masked out (setting their voxels to zero) from the testing phase’s input data (Fig. 4A). We evaluated the performance of the different models (i.e., with each occluded lobe), relative to the intact model, quantifying the percentage of change in the model’s accuracy with reference to the original results.

### Clustering based on neuropsychological assessments

Neuropsychological testing was comprised of six measures assessing three different cognitive domains: (1) Memory: Rey auditory verbal learning test (RAVLT) *(60)*, 30-minute delayed free recall and the RAVLT recognition test. (2) Language: Animal fluency *(55)* and the 30-item Boston naming test total score *(62)*. (3) Attention/Executive: Trail making test (TMT) *(63)*, part A and part B. Similar to previous research *(15)* raw scores in these neuropsychological measures were used to cluster MCI subjects into subgroups. Raw scores were first transformed into age- and education-adjusted z-scores based on means and standard deviations in each measure, calculated in the CN group. Then, an agglomerative hierarchical clustering analysis was performed on the z-scores using Ward’s method *(64)*. The clustering analysis resulted in four distinct subgroups *(15)*: *amnestic* MCI, *dysnomic* MCI, *dysexecutive* MCI, and *cluster-derived normal* group.

### Statistical analysis and visualization

All analyses were performed using the R statistical software, version 4.0.2 (https://www.r-project.org). Group differences in continuous variables were analyzed with an ANOVA or with independent-sample t-tests. ANOVAs were followed, when relevant, by Tukey post-hoc comparisons. Chi-squared tests were applied to evaluate differences in categorical variables. Longitudinal data were analyzed using the R WRS2 package *(65)*, and were based on a RM-ANOVA, with group (MCI-AD, MCI-CN) serving as the between-subjects factor and time (baseline, follow-up) as the within-subject factor. Mauchly’s test was applied to test for violations in the assumption of sphericity, followed by Greenhouse-Geisser corrections, if necessary. The plots for comparing cognitive scores, PET, and CSF between subgroups were based on the R RainCloudPlots package *(66)*. Regional imaging results were displayed on a surface using the R ggseg (https://lcbc-uio.github.io/ggseg/) package.

## Data Availability

All data associated with this study are in the paper or the Supplementary Materials. All raw data including MRI, CSF, and cognitive scores are available through the ADNI data archive (http://adni.loni.ucs.edu/).

## Acknowledgments

### Funding

Research reported in this publication was supported by the National Institute On Aging of the National Institutes of Health under Award Number R01AG062590. The content is solely the responsibility of the authors and does not necessarily represent the official views of the National Institutes of Health. Data collection and sharing for this project was funded by the Alzheimer’s Disease Neuroimaging Initiative (ADNI) (National Institutes of Health Grant U01 AG024904) and DOD ADNI (Department of Defense award number W81XWH-12-2-0012). ADNI is funded by the National Institute on Aging, the National Institute of Biomedical Imaging and Bioengineering, and through generous contributions from the following: AbbVie, Alzheimer’s Association; Alzheimer’s Drug Discovery Foundation; Araclon Biotech; BioClinica, Inc.; Biogen; Bristol-Myers Squibb Company; CereSpir, Inc.; Cogstate; Eisai Inc.; Elan Pharmaceuticals, Inc.; Eli Lilly and Company; EuroImmun; F. Hoffmann-La Roche Ltd and its affiliated company Genentech, Inc.; Fujirebio; GE Healthcare; IXICO Ltd.;Janssen Alzheimer Immunotherapy Research & Development, LLC.; Johnson & Johnson Pharmaceutical Research & Development LLC.; Lumosity; Lundbeck; Merck & Co., Inc.;Meso Scale Diagnostics, LLC.; NeuroRx Research; Neurotrack Technologies; Novartis Pharmaceuticals Corporation; Pfizer Inc.; Piramal Imaging; Servier; Takeda Pharmaceutical Company; and Transition Therapeutics. The Canadian Institutes of Health Research is providing funds to support ADNI clinical sites in Canada. Private sector contributions are facilitated by the Foundation for the National Institutes of Health (www.fnih.org). The grantee organization is the Northern California Institute for Research and Education, and the study is coordinated by the Alzheimer’s Therapeutic Research Institute at the University of Southern California. ADNI data are disseminated by the Laboratory for Neuro Imaging at the University of Southern California.

## Author contribution

K.K and E.D conceived research; K.K. analyzed data; K.K, K.S.G, M.S and E.D interpreted results. K.K and E.D. wrote the paper.

## Competing interest

The authors declare that they have no competing interests.

## Notes

### Competing Interest Statement

The authors have declared no competing interest.

### Author Declarations

The study involved analysis of de-identified data and was exempt from the local Institutional Review Board (UNC Chapel Hill, IRB number: 19-1090)

