## Supplementaly_information for "Atrophy-centered subtyping of mild cognitive impairment"

Eran Dayan, Ph.D.

Address: 130 Mason Farm Road, CB 7513, Chapel Hill, NC, 27599

^†^Data used in the preparation of this article were obtained from the Alzheimer’s Disease Neuroimaging Initiative (ADNI) database ([http://adni.loni.usc.edu](http://adni.loni.usc.edu/)). As such, the investigators within the ADNI contributed to the design and implementation of the ADNI and/or provided data but did not participate in analysis or writing of this article. A complete listing of ADNI investigators can be found at <http://adni.loni.usc.edu/wp-content/uploads/how_to_apply/ADNI_Acknowledgement_List.pdf>.

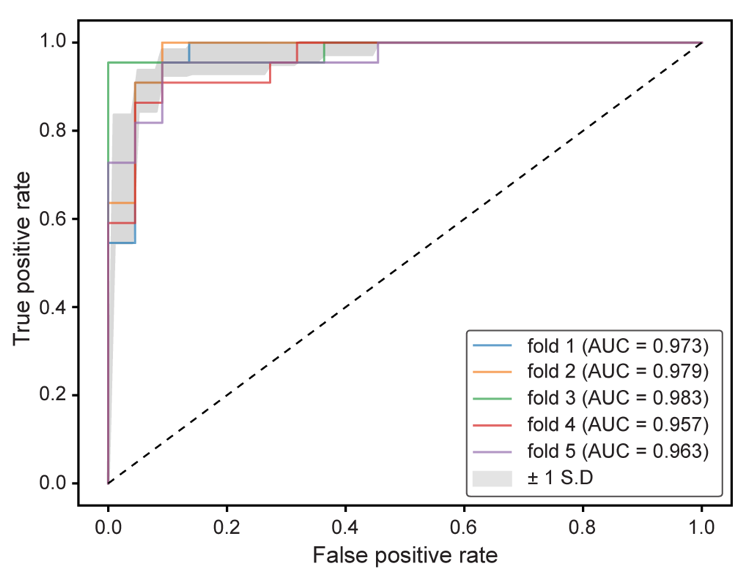

**Fig. S1. ROC curves for the proposed deep learning model obtained in each cross-validation fold.** Comparison of ROC curves obtained in each cross-validation fold revealed consistent performance of the proposed deep learning model during the training phase. Abbreviations: ROC=receiver operating characteristic, AUC=area under the curve, S. D=standard deviation.

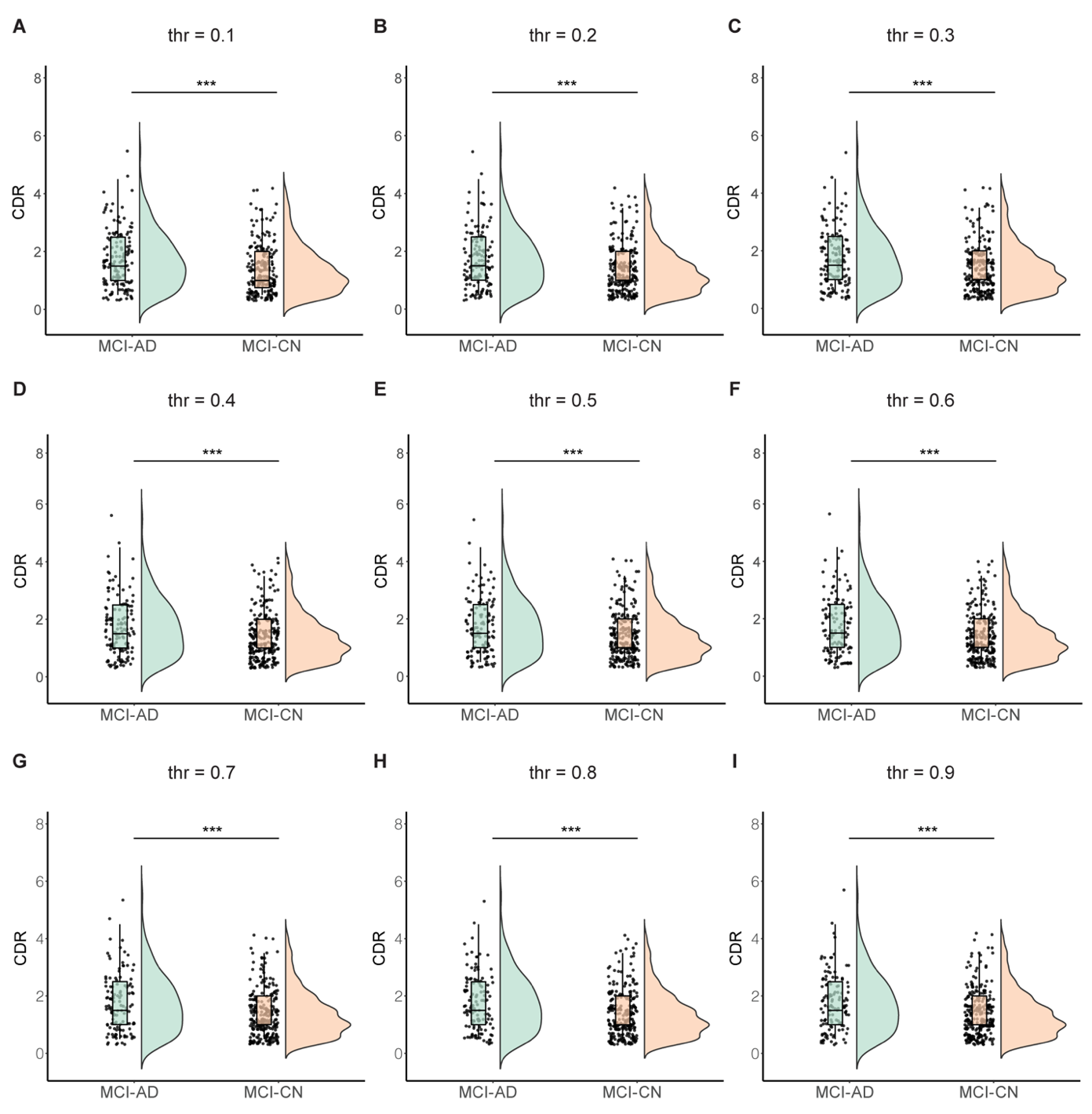

**Fig. S2. The effect of threshold criteria on MCI subgroup classification.** Threshold values used to assign class labels were manipulated, with the default threshold being 0.5 as the cut-off point. In each step, suprathreshold cases are assigned the label MCI-AD, while subthreshold cases are assigned the label MCI-CN. The manipulation of threshold (**A** to **I**) had relatively little effect on group differences, assessed here with CDR scores. Abbreviations: AD=Alzheimer’s disease, CN=cognitively normal, MCI=mild cognitive impairment, CDR=clinical dementia rating. ***p<0.001.

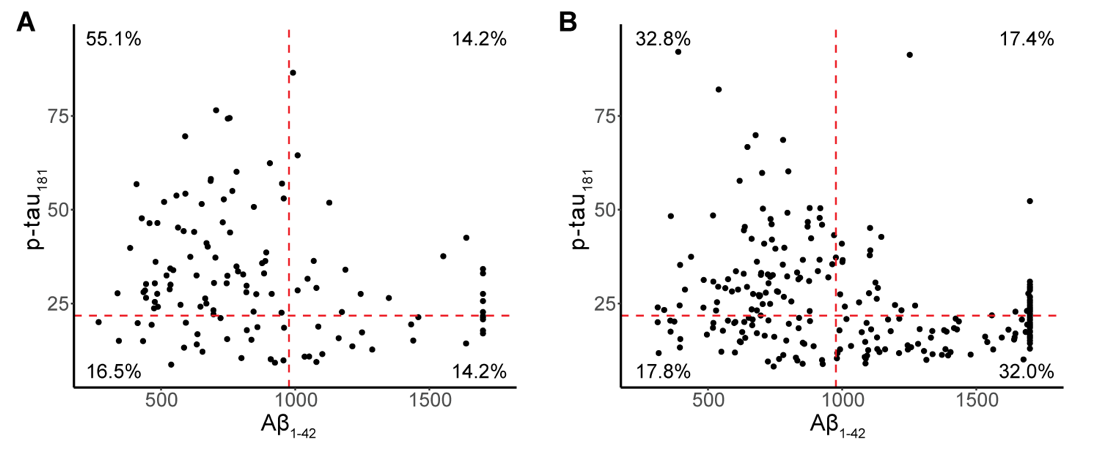

**Fig. S3. Distributions of CSF biomarker levels in the MCI subgroups.** The concordance between CSF A$\beta$_1-42_, and CSF p-tau_181_ levels are shown in the MCI-AD (**A**) and MCI-CN (**B**) subgroups. The red dashed lines indicate a-priori cut-off points used in the CSF biomarkers. The percentages indicate proportions of subjects falling in each quadrant. Abbreviations: AD=Alzheimer’s disease, CN=Cognitively normal, MCI=mild cognitive impairment, CSF= cerebrospinal fluid.

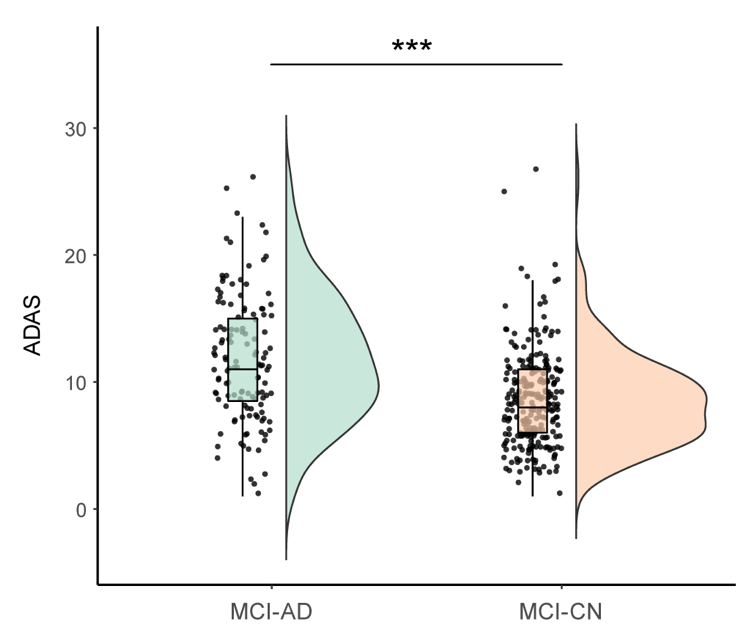

**Fig. S4.** **Comparison of cognition between the MCI subgroups.** Box plots show ADAS score in the MCI-AD and MCI-CN subgroups. Abbreviations: AD=Alzheimer’s disease, CN=Cognitively normal, MCI=mild cognitive impairment, ADAS=Alzheimer’s disease assessment scale. ****p*<0.001.

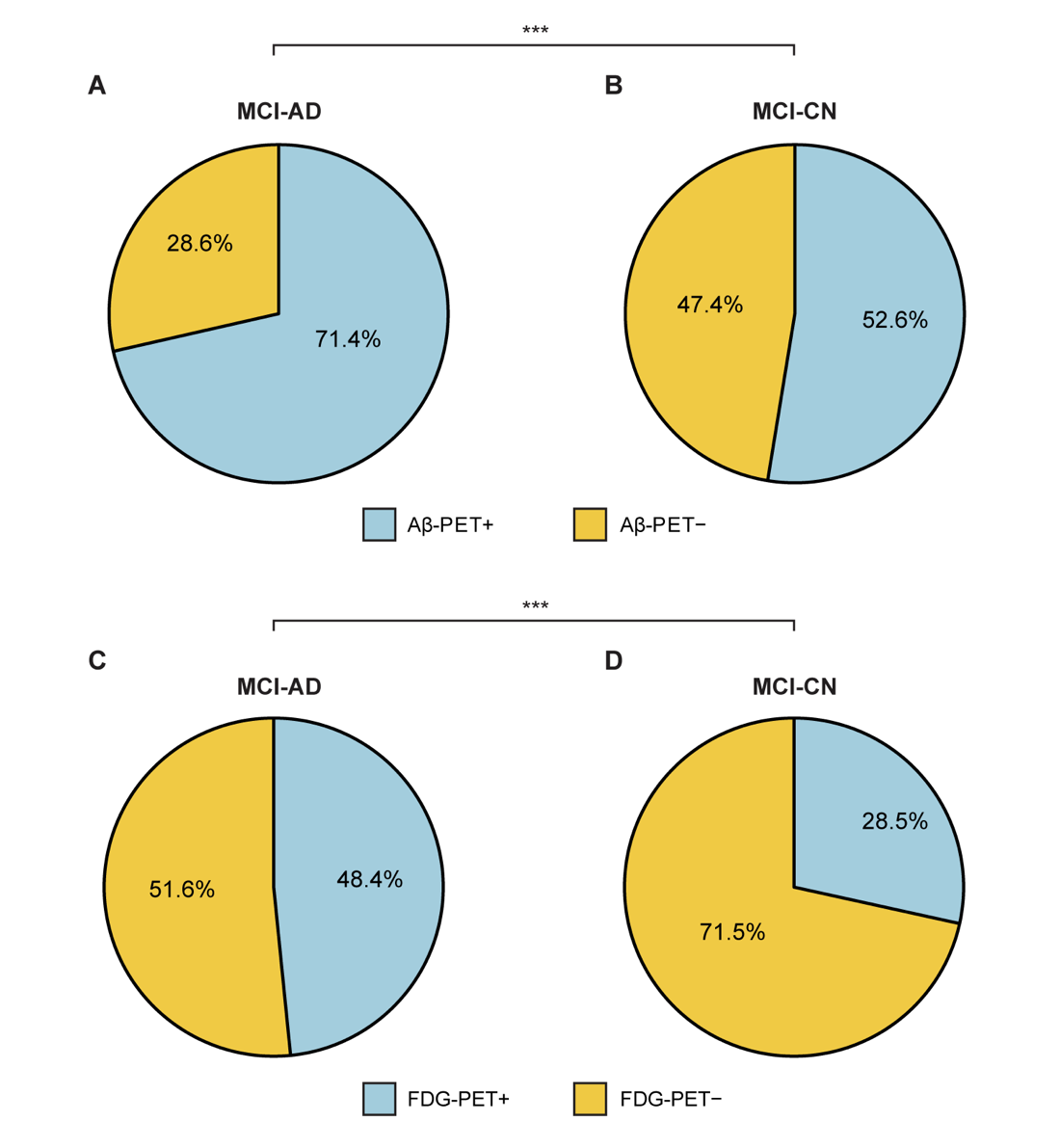

**Fig. S5. The percentage of normal/abnormal PET uptake in the MCI subgroups.** Pie charts depict the proportion of subjects in the MCI-AD (**A**) and MCI-CN (**B**) subgroups with normal and abnormal A$\beta$-PET; The proportion of subjects in the MCI-AD (**C**) and MCI-CN (**D**) subgroups with normal and abnormal FDG-PET. The distributions of normal and abnormal PET uptake values were significantly different between the two subgroups (A$\beta$-PET: $\chi^{2}$ = 12.35, *p* < 0.001; FDG-PET: $\chi^{2}$ = 14.70, *p* < 0.001). Abbreviations: AD=Alzheimer’s disease, CN=Cognitively normal, MCI=mild cognitive impairment, A$\beta$-PET =beta-amyloid positron emission tomography, FDG-PET =fluorodeoxyglucose positron emission tomography. ****p*<0.001.

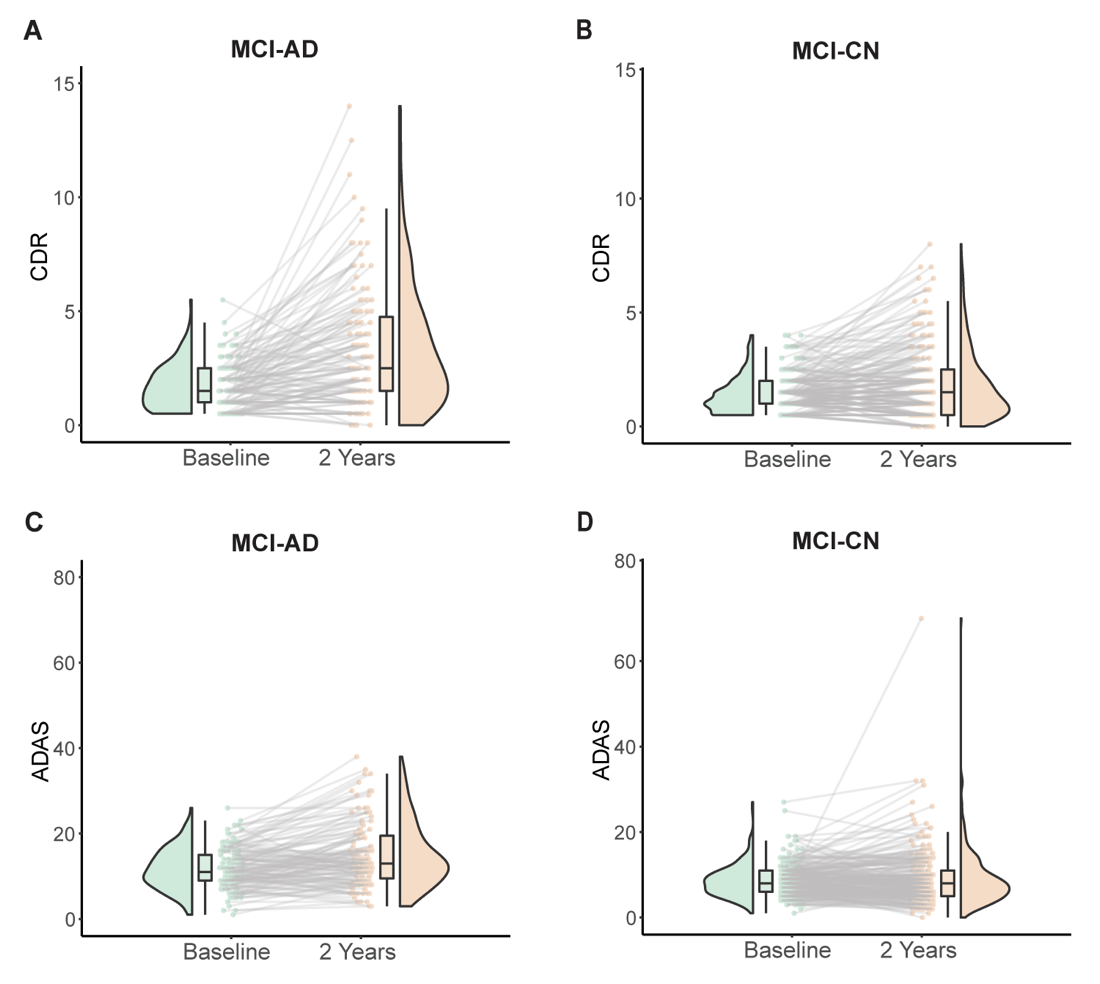

**Fig. S6. Longitudinal changes in cognition and clinical rating observed in the MCI subgroups**. The plots show longitudinal changes in cognition and clinical rating scores, observed at baseline and at the 2^nd^ year follow-up**.** Longitudinal changes are show for: CDR scores (**A,B**) and ADAS scores (**C,D**). Abbreviations: MCI=mild cognitive impairment, CDR=clinical dementia rating, ADAS=Alzheimer’s disease assessment scale.

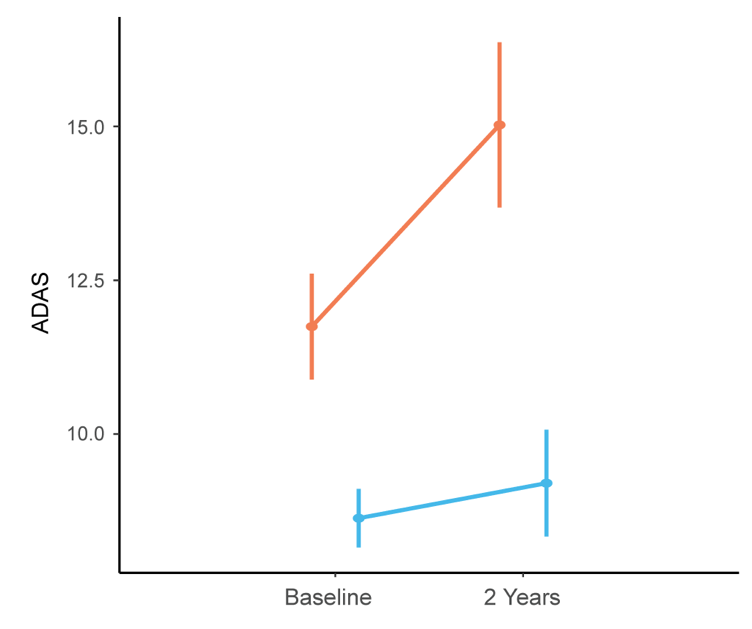

**Fig. S7. Longitudinal comparison of cognition in the MCI subgroups.** Longitudinal changes in cognition in the 2 MCI subgroups were tested with a RM-ANOVA. Comparisons focused on ADAS scores. Abbreviations: MCI=mild cognitive impairment, ADAS=Alzheimer’s disease assessment scale, RM-ANOVA=repeated measures-analysis of variance.

**Table S1. Group comparisons.**

| **Variable** | **Group** | **Mean difference** | **95 % CI** | | **P value** |
| --- | --- | --- | --- | --- | --- |
|  |  |  | **Lower Bound** | **Upper Bound** |  |
| CDR | CN (A−T−) vs AD (A+T+)  MCI vs AD (A+T+)  MCI vs CN (A−T−) | -4.57  -3.08  1.48 | -4.89  -3.34  1.22 | -4.24  -2.82  1.75 | < 0.001  <0.001  <0.001 |
| ADAS | CN (A−T−) vs AD (A+T+)  MCI vs AD (A+T+)  MCI vs CN (A−T−) | -15.00  -12.34  2.67 | -16.55  -13.56  1.43 | -13.47  -11.11  3.91 | <0.001  <0.001  <0.001 |
| CSF A$\beta$_42_ | CN (A−T−) vs AD (A+T+)  MCI vs AD (A+T+)  MCI vs CN (A−T−) | 856.96  389.43  -467.53 | 741.99  297.33  -559.96 | 971.93  481.54  -375.10 | <0.001  <0.001  <0.001 |
| CSF p-tau_181_ | CN (A−T−) vs AD (A+T+)  MCI vs AD (A+T+)  MCI vs CN (A−T−) | -24.02  -12.82  11.20 | -28.32  -16.27  7.74 | -19.71  -9.37  14.66 | <0.001  <0.001  <0.001 |

Post-hoc comparisons using Tukey’s HSD. Abbreviations: CI=Confidence interval, AD=Alzheimer’s disease, CN=cognitively normal, MCI=mild cognitive impairment, CDR=clinical dementia rating, ADAS=Alzheimer’s disease assessment scale, A$\beta$_42_=beta-amyloid42, p-tau_181_=phosphorylated-tau181.

**Table S2. Demographics of subtype based on neuropsychological assessments.**

|  | **Dysnomic MCI** | **aMCI** | **Dysexecutive MCI** | **Clustered-Derived Normal** |
| --- | --- | --- | --- | --- |
| N | 140 | 137 | 26 | 71 |
| **Memory** |  |  |  |  |
| RAVLT delayed recall | 3.6 (3.2) | 2.3 (2.1) | 3.1 (2.9) | 9.7 (3.0) |
| RAVLT recognition | 11.0 (3.4) | 9.8 (2.8) | 9.6 (2.4) | 14.2 (1.1) |
| **Attention-Executive function** |  |  |  |  |
| TMT part A | 46.5 (9.0) | 30.3 (6.1) | 81.6 (23.3) | 28.1 (6.4) |
| TMT part B | 122.6 (57.8) | 93 (44.3) | 197.7 (72.2) | 81.3 (41.8) |
| **Language** |  |  |  |  |
| Boston naming test | 25.2 (4.5) | 27.9 (1.7) | 24.0 (4.2) | 28.1 (1.5) |
| Animals fluency | 15.6 (4.1) | 18.6 (4.0) | 12.1 (4.0) | 22.8 (4.5) |

Continuous variables are presented as means with SDs. Abbreviations: MCI=mild cognitive impairment. N=number of subjects, RAVLT= Rey auditory verbal learning test, TMT=trail making test, SD=standard deviation.
